# Software application profile: MeltingPlot, a user-friendly online tool for epidemiological investigation using High Resolution Melting data

**DOI:** 10.1101/2020.06.16.20132142

**Authors:** Matteo Perini, Gherard Batisti Biffignandi, Domenico Di Carlo, Ajay Ratan Pasala, Aurora Piazza, Simona Panelli, Gian Vincenzo Zuccotti, Francesco Comandatore

**Author notes:** Corresponding author., postal: Pediatric Clinical Research Center “Romeo and Enrica Invernizzi”, Via Giovanni Battista Grassi, 74, 20157 Milano, Italy.

## Abstract

**Motivation:** Genomic investigations show that nosocomial outbreaks can be sustained by the simultaneous spreading of multiple pathogen clones. Rapid pathogen typing and real-time clones monitoring is a key strategy to control pathogen spreading in hospital settings. A novel approach to High Resolution Melting (HRM) data analysis allows pathogen typing in less than 5 hours. MeltingPlot is the first tool specifically designed for epidemiological investigations using HRM data. The tool is suitable for large real-time surveillance and rapid outbreak reconstructions.

**Implementation:** MeltingPlot was developed in R. The web tool and the standalone versions are available.

**General features:** The tool implements a graph-based algorithm designed to discriminate pathogen clones on the basis of HRM data, producing portable typing results. MeltingPlot also merges typing information with isolates and patients metadata to create graphical and tabular outputs useful in epidemiological investigations.

**Availability:** The web interface is available at https://skynet.unimi.it/index.php/tools/meltingplot, the standalone version at https://github.com/MatteoPS/MeltingPlot.

## Introduction

The rapid typing of pathogens is pivotal to perform fast epidemiological investigations and to detect and block outbreaks. High-Resolution Melting (HRM) analysis is a single-step molecular biology technique able to discriminate sequence variants measuring the melting temperature of PCR amplicons. This allows pathogen typing in less than 5 hours [1,2]. For each isolate, HRM analysis interrogates *n* specific genomic regions returning *n* melting temperatures, where each genomic region is defined by a specific PCR primer set. The melting temperatures of each interrogated genomic region depend on its nucleotide composition. Consequently, melting temperatures can be used to cluster the isolates in a *n*-dimensional space. Previously, we developed a graph-based algorithm for isolate clustering on the basis of HRM temperatures, and we successfully validated this approach on 82 isolates of *Klebsiella pneumoniae* [3], one of the most important nosocomial pathogens world-wide [4].

Here we present MeltingPlot, a tool for rapid epidemiological investigation using HRM data. The tool implements an evolution of the clustering algorithm we already published [3]. Moreover, MeltingPlot merges HRM typing information with metadata of isolates and patients to get a comprehensive epidemiological investigation. MeltingPlot has a user-friendly web interface (the standalone command line version is also available) and it creates easy to read graphical and tabular outputs. The tool runs in a few seconds even with hundreds of isolates.

### Implementation

The flow of MeltingPlot can be divided in three main steps: HRM-based clustering/typing of isolates, prevalence analysis and transmission analysis. HRM-based clustering/typing is computed on the basis of the High Resolution Melting (HRM) temperatures of the isolates amplicons, these temperatures are the only input needed for this step. After the computation of the average HRM temperature of the technical replicates, the isolates are organized in a graph where the vertices are the isolates and two vertices are connected if the difference of their average HRM temperatures is less or equal to 0.5 °C for each PCR primer set used in the HRM typing method. The graph is then decomposed into separate components (groups of connected vertices) and each one is then divided in clusters using the Edge Betweenness Clustering algorithm [5] implemented in the cluster_edge_betweenness function of the igraph R library [6]. Briefly, the betweenness centrality of each edge of the graph was computed as the number of shortest paths that go through the edge, and clusters were identified by gradually removing the edges with the highest betweenness centrality values. High betweenness centrality values among two vertices indicates that the two vertices most probably do not belong to the same cluster and *vice versa*.

Furthermore, the betweenness centrality of each vertex of the graph was computed as the number of graph short paths passing that vertex. Vertex with higher betweenness centrality values are those that connect two or more clusters. We used this parameter to identify vertices that were not strongly associated with a single cluster. Thus, vertices with normalized betweenness centrality values above a threshold were not assigned to any cluster and they were classified as “undetermined” by the tool (this threshold of normalized betweenness value can be set by the user, the default is 0.5).

Unfortunately, HRM-based clustering results obtained from different datasets are not directly comparable. To obtain comparable HRM typing results, the user can include in the analysis the HRM temperatures of a collection of reference strains: isolates previously analysed by the same HRM protocol and for which typing annotation is known (e.g. Sequence Type). When a reference collection is provided, MeltingPlot labels each cluster with the annotation of the reference isolates contained in it. For details see the Supplementary Material.

Prevalence analysis and transmission analysis steps can be performed only when patients/isolates metadata is provided. In these steps the tool joins the HRM clustering results with the isolates metadata to create various outputs that depict the spreading of pathogen clones among wards and patients over time. For more details see the output files section below or the Supplementary Material. MeltingPlot was developed in R and its dependencies are the libraries igraph [6], gplots [7], xlsx [8], ggplot2 [9], scales [10]. The user interface on the website was developed in PHP.

### Use

#### Input file

Users are required to download and fill an xls template spreadsheet that contains four sheets: *HRM_temperatures, Isolates_metadata, Reference_isolates* and an *HELP_notes* sheet:

- *HRM_temperatures:* in this sheet the user has to report the HRM temperatures of the study isolates. This is the only mandatory data and it is used to perform the HRM-based clustering/typing analysis. If HRM experiments were performed using technical replicates, the users have to report all the replicate temperatures;
- *Isolates_metadata*: in this sheet the users can provide patients/isolates metadata, e.g. isolation date, isolation location (e.g. hospital ward) and an ID for the patients (e.g. Pz1, Pz2, …). This information is not mandatory for HRM isolates typing but it is required to perform the complete epidemiological investigation (i.e. prevalence analysis and transmission analysis);
- *Reference_isolates*: this sheet contains the HRM temperatures of the reference isolates and their annotation (e.g. Sequence Type). The reference isolates annotation will be used to label the clusters, making the obtained HRM typing results portable when the same reference collection is used.
- *HELP_notes*: this sheet contains important information about the rules for each column of the spreadsheet.

All the templates (the blank template, the templates with reference HRM temperature collections, and the example files) are available on the MeltingPlot webpage.

#### Output files

MeltingPlot creates three groups of plot files (in PDF and PNG format), one for each step of the analysis: the HRM-based clustering/typing, the prevalence analysis and the transmission analysis. The HRM-based clustering/typing plot group includes the isolates graph (where each isolate is colored on the basis of its cluster) and a heatmap showing the HRM temperatures and the isolates clusters. In the isolates graph each vertex is an isolate and two isolates are connected as described above. The last two groups are created when isolates metadata is provided. The prevalence analysis plot group includes bar plots showing the distribution of the clusters over time in the different locations. The transmission analysis plot group contains a patient timeline and a patient-to-patient graph. In this graph, two patients are connected when two isolates belonging to the same HRM cluster were collected from both patients. The edge is thicker when the isolates were collected in the same location (e.g. ward) within a number of days set by the user (7 by default). Thus, thicker edges highlight most probable transmission events. MeltingPlot also produces xls spreadsheets containing the isolates HRM clusters and metadata. See Supplementary Material for details.

### Example of epidemiological investigation

To show the capability of MeltingPlot, we simulated a large *K. pneumoniae* outbreak (100 isolates) sustained by multiple clones, a situation observed in real nosocomial outbreaks [11]. We used HRM temperatures extracted from a dataset of *K. pneumoniae* isolates previously analyzed in our laboratory. We included in the analysis a reference collection of 18 representative isolates out of the 82 previously typed by HRM and WGS [3] (this temperature collection is available on the tool web site). In a real hospital setting, the HRM typing of the 100 isolates would be performed after every pathogen isolation during the entire outbreak period (∼3 months). The entire real-time epidemiological investigation would cost ∼500 euros. As shown in Fig. 1, the outbreak is sustained by three major isolate clusters. MeltingPlot labelled these clusters as *wzi173_(ST307)* (in red), *wzi154_(ST512/ST258)* (in green) and *wzi89_(ST15)* (in violet) using the annotation of the reference isolates. The main outbreak is caused by the two pathogen clusters (green and red). Each of these clusters is highly associated with a single ward: the green one with Ward A and the red one with Ward B. The violet cluster causes a smaller outbreak in Ward C at the beginning of the investigated period. The patient’s timeline and the patient-to-patient graph clearly show that two patients (Pz15 and Pz17) were infected by isolates of the red and green clusters and they also crossed the wards A and B: this highlights two possible pathogen transmission routes among the wards. A complete description of each output file is available in the Supplementary Material.

**Fig. 1.**
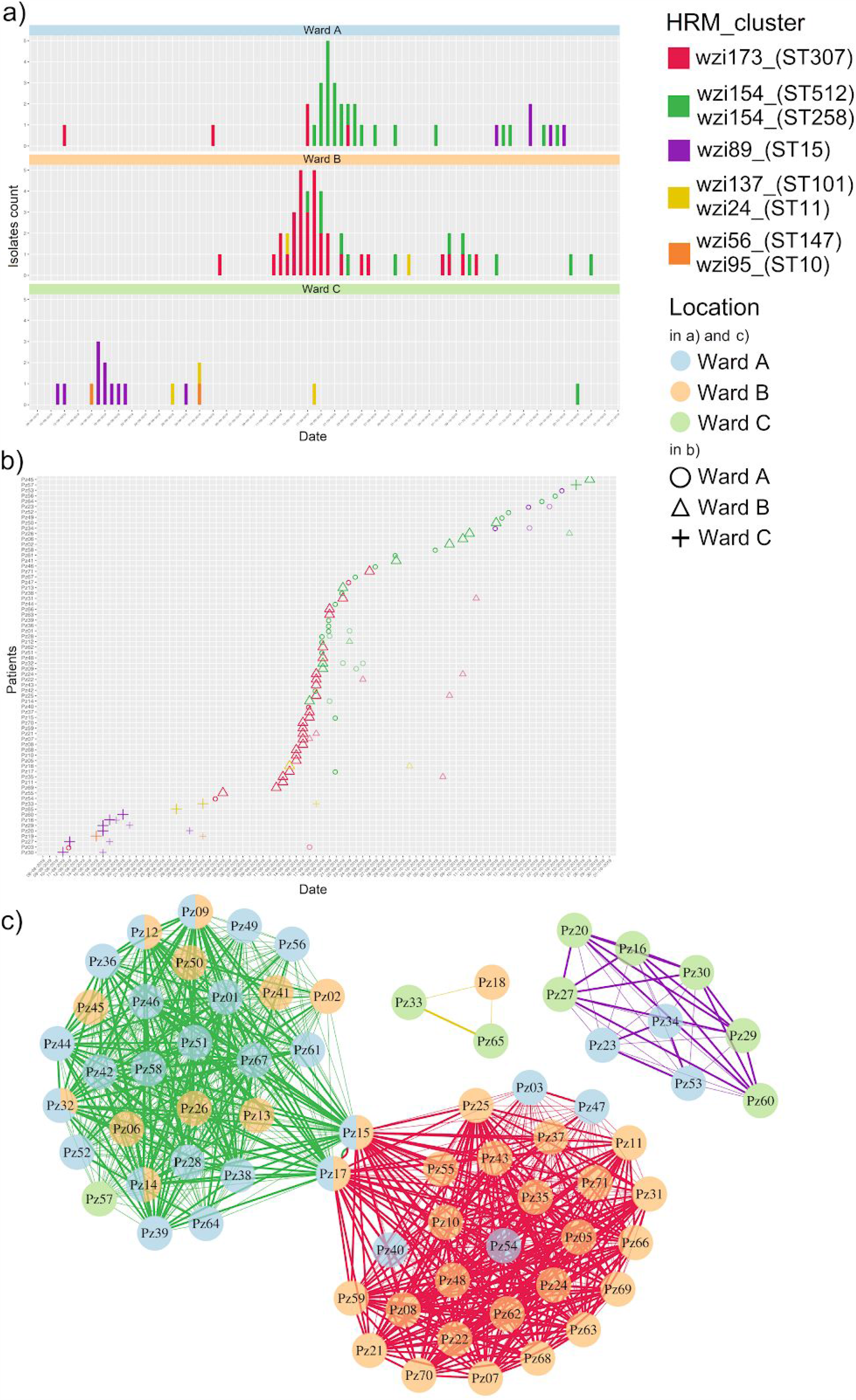
MeltingPlot Output, example of an epidemiological investigation. Here we report the three most significant MeltingPlot output plots obtained on the simulated dataset. The plots were selected to show the power of the HRM-based epidemiological investigation performed by the tool. Higher resolution images are available in the Supplementary Material. **a) Prevalence analysis**: the plot shows the number of isolates collected from each hospital ward over time. Each HRM cluster is represented with a different color. This analysis allows the detection of the pathogen clones emergence in the hospital setting. **b) Patients’ timeline**: each row refers to a patient and the symbols represent isolates. The shape of the symbols report the location where the isolates were collected while the colors indicate the HRM cluster. **c) Patient-to-patient graph**: each vertex represents a patient and two vertices are connected if isolates belonging to the same HRM cluster were collected from both patients. Vertices are reported as pie charts and colors show the locations (wards) where the isolates of the patients were collected. The edges of the graph are thicker if the isolates from the same HRM cluster were collected within seven days (this threshold can be defined by the user) from the same location. This plot can help to identify the transmission routes of the pathogen in the hospital setting.

## Discussion

High Resolution Melting (HRM) is a fast and inexpensive molecular biology technique [2] applicable to pathogen typing and suitable for large scale surveillance programmes as well as for fast outbreak reconstruction [1]. Recently, we proposed a novel approach to HRM-based pathogen typing based on the idea that HRM temperatures can be used to cluster genetically similar strains [3]. We validated this approach on the nosocomial bacterial pathogen *Klebsiella pneumoniae* [3]. In this work we propose MeltingPlot, a tool that implements an evolution of the HRM-based clustering algorithm we already used [3]. The tool allows also to perform epidemiological investigation and transmission analysis using HRM data.

HRM-based clustering analysis groups isolates that show similar melting temperatures. Unfortunately, the results of HRM-based clustering analysis on different collections of isolates can be difficult to compare. To overcome this limitation we made MeltingPlot able to include in the clustering analysis the HRM temperatures of a collection of reference isolates and to use them to annotate the obtained clusters and the study isolates.

MeltingPlot performs complete epidemiological investigations merging HRM clustering results with isolates/patients metadata. It produces easy-to-read graphical representations and tabular files. As shown in the example of epidemiological investigation (see above) MeltingPlot is useful to reconstruct epidemiological scenarios and to identify pathogen transmission routes. Furthermore, the web interface makes the tool user-friendly and the user has only to upload the data into an xls template spreadsheet. The MeltingPlot analyses hundreds of isolates in a few seconds.

HRM technique allows pathogen typing in a few hours and ∼5 euros per sample. Despite this, the mathematical/informatic skills required for the analysis and interpretation of HRM results limit the application of HRM typing protocols in hospital real time surveillance. MeltingPlot is a user-friendly tool that facilitates the application of HRM to real time large scale surveillance programs in hospital settings.

## Data Availability

All data used in the manuscript are reported at https://skynet.unimi.it/wp-content/uploads/MeltingPlot/files_supplementary_material/big_example.xls

## Acknowledgements

Thanks to the Romeo ed Enrica Invernizzi Foundation.

